# Pre-pregnancy primary care and accident and emergency interactions among interpreter-users versus non-interpreter-users: a retrospective cohort study

**DOI:** 10.1101/2025.07.03.25330826

**Authors:** Majel McGranahan, Nuria Sanchez Clemente, Lars Murdock, Baboucarr Njie, Yamina Boukari, Zach Welshman, Alexia Sampri, Oyinlola Oyebode, Felicity Boardman, Robert Aldridge, Neha Pathak, the CVD-COVID-UK/COVID-IMPACT Consortium

## Abstract

**Introduction:** Non-English-speaking migrant women face healthcare barriers, exacerbating maternal health inequalities. Primary care-based preconception interventions can reduce preconception risk factors but require engagement. We explored whether women interact with general practice (GP) and accident and emergency (A&E) in the year pre-pregnancy, differences between interpreter-users and non-interpreter-users, and changes over the COVID-19 pandemic.

**Methods:** English population-wide linked data including primary, secondary and maternity-care were used. Participants included women with estimated pregnancy start dates 1/3/2019-29/2/2024, aged 18-49.

Outcome measures were interactions with GP and A&E in the year pre-pregnancy (yes/no) according to interpreter-use. Outcomes were recorded according to pregnancy start date: 1/3/2019-29/2/2020 (pre-pregnancy-year pre-COVID-19-onset), 1/3/2020-28/2/2021 (pre-pregnancy-year overlapping with COVID-19-onset) or 1/3/2020-29/2/2024 (pre-pregnancy-year after COVID-19-onset). Logistic regression compared GP/A&E interaction pre-pregnancy among interpreter-users versus non-interpreter-users.

**Results:** Among 2,182,280 women, 61,140 (2.8%) used interpreters. Median age was 31.0 among interpreter-users and 30.9 among non-interpreter-users. 49.7% (n=30,370) of interpreter-users were in the most deprived quintile versus 22.7% (n=480,470) of non-interpreter-users, and 62.5% (n=38,150) were of ethnic minorities (excluding white minorities) versus 22.1% (n=468,530) of non-interpreter-users.

79.5% (n=48,625) of interpreter-users interacted with GP in the year pre-pregnancy, versus 85.0% (n=1,802,925) of non-interpreter-users. Interpreter-use was associated with lower adjusted odds (aOR) of GP interaction, including among women whose pre-pregnancy-ear was pre-COVID-19-onset (aOR 0.92 [95%CI 0.88-0.96]), overlapped with COVID-19-onset (aOR 0.76 [95%CI 0.73-0.79]) and post-COVID-19-onset (aOR 0.63 [95%CI 0.61-0.65]).

Adjusted odds of A&E interaction in the year pre-pregnancy were 0.96 (95%CI 0.95-0.98) lower among interpreter-users. Odds of A&E interactions did not change substantially during COVID-19.

**Conclusions:** GP interactions may improve preconception health. Ensuring interpreter-need is recorded at GP registration is important to ensure communication in appropriate languages. Widening inequalities in GP interactions suggest a proactive approach may be required.

**Key Messages:** Non-English-speaking women face healthcare barriers, and primary care-based preconception interventions can reduce preconception risk factors. However, previously, we did not know whether women using interpreters interact with GP or A&E differently from non-interpreter-users, and whether and how interactions with GP/A&E pre-pregnancy have changed since COVID-19. This research shows that most women interact with the GP at some point in the year pre-pregnancy, but fewer interpreter-users do so, and inequalities have increased since COVID-19 suggesting a more proactive approach to preconception health may be needed for this group. Most women do not interact with A&E in the year before getting pregnant. Therefore, clinicians in primary care are ideally placed to offer preconception health interventions but targeted interventions in multiple languages may be needed outside GP for those not accessing GP care before they get pregnant.

## Introduction

In 2023, nearly one in three UK births were to women* born abroad^1^, many of whom require interpreters when accessing healthcare^2^. Maternal health inequalities exist between migrant and non-migrant women^3^ and a woman’s preconception health has a strong influence on pregnancy outcomes^4^. Analysis of the UK’s national Maternity Services Dataset showed that migrant women in vulnerable situations are less likely to take folic acid pre-pregnancy, more likely to have hepatitis B and diabetes preconception, and to access antenatal care late compared with UK-born women ^5^. Non-English-speaking women face additional barriers to care^6^.

Primary care-based preconception interventions, such as intensive or brief education, diet modification and supplementary medication can reduce preconception risk factors^7^. Although no previous studies could be identified on preconception interventions in an Accident and Emergency (A&E) setting, A&E could provide an opportunity to intervene preconception for those who are less likely to access general practice (GP).

A UK cross-sectional study of primary care records of 193,578 women from 2017-2018 found that 86.6% of women had face-to-face contact with a GP or nurse during the year before pregnancy^8^. Although primary care appointment volumes decreased during COVID-19 lockdowns, they have since recovered to pre-pandemic levels for the general population^9^. However, little is known about whether women using interpreters access care differently, how COVID-19 impacted GP interactions preconception and, if women interact with GP differently preconception since COVID-19, whether any differences vary by interpreter-use.

Moreover, although deprivation has been associated with increased A&E attendance in general^10^, no previous study has explored A&E attendance in the year preconception, or whether there are differences in attendance between interpreter-users and non-interpreter-users, so it is unknown whether A&E could be another avenue for preconception health improvement interventions.

To develop effective opportunistic interventions to improve preconception health within GP or A&E, we need an up-to-date understanding of how women interact with GP and A&E before getting pregnant and whether this has changed with COVID-19. We also need an understanding of whether interpreter-users interact differently with GP and A&E, so that interventions can be adapted if necessary.

We aimed to describe whether women interact with GP and A&E in the year pre-pregnancy, differences between interpreter-users and non-interpreter-users, and changes before, during and after the onset of COVID-19.

## Materials and Methods

### Study design

This retrospective observational cohort study used data from an English population-based national person-level linked dataset of primary care (General Practice Extraction Service Data for Pandemic Planning and Research, GDPPR^11^), secondary care (Hospital Episode Statistics) and maternity records (Maternity Services Data Set – MSDS) within NHS England Secure Data Environment (SDE) service for England, accessed through the British Heart Foundation (BHF) Data Science Centre’s CVD-COVID-UK/COVID-IMPACT Consortium. The methods for data linkage have been described in detail elsewhere^12^.

### Study population

Participants included women with singleton pregnancies recorded within MSDS with estimated pregnancy start dates between 1/3/2019-29/2/2024, aged 18-49 years at estimated pregnancy start date, who were alive on 1/11/2019, with a GP record at least one year before the estimated pregnancy start date at one of the 98% of participating GPs in England ^13^ (see Supplementary File 1 for how estimated pregnancy start date was calculated). For women with multiple pregnancies during the study period, only their first pregnancy during the study period (regardless of pregnancy outcome) was included. We excluded women who had a preceding pregnancy before the start of the study period if it was within the year pre-pregnancy of an index pregnancy (as it was not possible to tell which pregnancy GP interactions related to).

### Exposure

SNOMED codes describing interpreter use within GP (see https://github.com/BHFDSC/CCU063_03/tree/main/phenotypes) were used to identify those who had used interpreters within GP at any point since the start of their record (the earliest recorded interpreter code was from 1974) until 4/2/2025. SNOMED codes are usually added to a patient’s record when there is some kind of interaction (or when the patient registers with a GP practice) in order to record events during that interaction. We adapted and added to a pre-existing codelist related to migrant codes^14^ to create the interpreter codelist.

### Outcomes

Outcome measures were interactions with GP (excluding prescriptions) and A&E attendances (including walk-in centres and minor injury units) in the year pre-pregnancy (yes/no), according to interpreter use (see https://digital.nhs.uk/coronavirus/gpes-data-for-pandemic-planning-and-research/guide-for-analysts-and-users-of-the-data#code-clusters-and-content for codes available within GDPPR). We also examined interactions with GP and A&E according to pre-pregnancy year relative to COVID-19 (before, overlapping with, or after 1/3/2020).

The 500 most frequently occurring codes were reviewed with a practising GP to ensure they corresponded to GP interactions. Codes not considered representative of GP interactions were removed (see Supplementary File 1). See Supplementary File 2 for the 500 most frequently occurring codes after removal of those not considered representative of GP interactions.

To understand the potential for selection bias, for participants with interpreter codes within their GP record, we summarised when an interpreter code was first recorded in relation to their pregnancy (before the year pre-pregnancy, during the year pre-pregnancy, during pregnancy, or after the recorded delivery date (or last recorded maternity interaction if delivery date unavailable)) to determine if it was possible that recorded interactions were due to interpreter-use being recorded during the pre-pregnancy year.

### Statistical analysis

Participants’ age (years), deprivation quintile (based on Index of Multiple Deprivation – an area-based relative deprivation measure), ethnic group (five categories - see Table 1), geographical region (nine categories – see Table 1), previous pregnancy (recorded from MSDS antenatal booking data, see Supplementary File 1 for details), and history of interpreter use in primary care were summarised using descriptive statistics. Age, deprivation quintile, geographical region and ethnicity were recorded according to BHF data science centre methods described elsewhere^15^ (code available from https://github.com/BHFDSC/hds_curated_assets/blob/main/D07-demographics.py). Where age was unavailable from the aforementioned BHF data science centre methods, the age derived from MSDS booking data was used.

**Table 1:** Demographic characteristics of study population (women with estimated pregnancy start dates between 1/3/2019-29/2/2024, aged 18-49)

|  | **Overall** | **Yes** | **No** |
| --- | --- | --- | --- |
| **Characteristic** | **(N=2,182,280)** | **(N=61,140)** | **(N=2,121,140)** |
| **Age (years)** |  |  |  |
| Mean (SD) | 30.7 (5.51) | 30.9 (5.84) | 30.7 (5.50) |
| Median (interquartile range) | 30.9 [23.3, 38.5] | 31.0 [22.5, 39.4] | 30.9 [23.3, 38.4] |
| **Ethnic group** |  |  |  |
| White | 1,673,735 (76.7%) | 22,930 (37.5%) | 1,650,805 (77.8%) |
| Black, Black British, Caribbean or African | 104,095 (4.8%) | 6,705 (11.0%) | 97,390 (4.6%) |
| Asian or Asian British | 286,355 (13.1%) | 20,780 (34.0%) | 265,575 (12.5%) |
| Mixed or multiple ethnic groups | 57,290 (2.6%) | 2,380 (3.9%) | 54,910 (2.6%) |
| Other ethnic group | 58,935 (2.7%) | 8,280 (13.5%) | 50,655 (2.4%) |
| Missing | 1,870 (0.1%) | 60 (0.1%) | 1,810 (0.1%) |
| **Deprivation quintile (Index of Multiple Deprivation)** |  |  |  |
| 1 (most deprived) | 510,840 (23.4%) | 30,370 (49.7%) | 480,470 (22.7%) |
| 2 | 457,555 (21.0%) | 16,480 (27.0%) | 441,075 (20.8%) |
| 3 | 426,940 (19.6%) | 7,690 (12.6%) | 419,250 (19.8%) |
| 4 | 406,720 (18.6%) | 4,165 (6.8%) | 402,560 (19.0%) |
| 5 (least deprived) | 378,015 (17.3%) | 2,420 (4.0%) | 375,600 (17.7%) |
| Missing | 2,205 (0.1%) | 15 (0.0%) | 2,190 (0.1%) |
| **Previous pregnancy** |  |  |  |
| No | 790,275 (36.2%) | 13,150 (21.5%) | 777,125 (36.6%) |
| Yes | 1,213,280 (55.6%) | 42,195 (69.0%) | 1,171,090 (55.2%) |
| Missing | 178,725 (8.2%) | 5,795 (9.5%) | 172,930 (8.2%) |
| **Region** |  |  |  |
| London | 359,525 (16 %) | 19,650 (32.1%) | 339,875 (16.0%) |
| South East | 356,430 (16.5%) | 3,135 (5.1%) | 353,290 (16.7%) |
| South West | 195,465 (9.0%) | 1,460 (2.4%) | 194,010 (9.1%) |
| East of England | 248,910 (11.4%) | 4,755 (7.8%) | 244,155 (11.5%) |
| East Midlands | 184,570 (8.5%) | 5,725 (9.4%) | 178,840 (8.4%) |
| West Midlands | 234,740 (10.8%) | 5,935 (9.7%) | 228,805 (10.8%) |
| Yorkshire and The Humber | 212,920 (9.8%) | 11,005 (18.0%) | 201,910 (9.5%) |
| North East | 98,405 (4.5%) | 2,225 (3.6%) | 96,180 (4.5%) |
| North West | 289,110 (13.2%) | 7,225 (11.8%) | 281,890 (13.3%) |
| Missing | 2,205 (0.1%) | 15 (0.0%) | 2,190 (0.1%) |

Two logistic regression models were developed to quantify the association between interpreter-use, the key exposure of interest, and a) GP interactions in the year pre-pregnancy (yes/no); and b) A&E interactions in the year pre-pregnancy (yes/no). Adjustment for confounding factors was based on factors available in the dataset that are known to be associated with healthcare attendance and likely to be associated with interpreter use.

Forced entry model fitting was used (covariates were added to the regression model simultaneously). Confounding factors adjusted for included mother’s age (years), deprivation quintile, ethnic group (five categories, see Table 1), geographical region (nine categories, see Table 1) and previous pregnancy (yes/no – according to MSDS antenatal booking data). For logistic regression analyses, participants with missing data for any covariate were excluded from the analyses.

Logistic regression analyses were undertaken for the whole study period, and according to whether participant’s pre-pregnancy year was before, overlapping with or after 1/3/2020.

To comply with NHS England statistical disclosure control guidance, counts were rounded to the nearest five and cells smaller than ten were supressed.

Analysis was undertaking following a prespecified protocol using R version 4.1.3, and data curation undertaken using Pyspark version 3.3.0 within Databricks. The study protocol and code used for the data curation and analysis are available from https://github.com/BHFDSC/CCU063_03.

### Sensitivity analysis

Because of the complex relationship between ethnicity and migration^16^, and therefore interpreter use, sensitivity analysis explored the contribution of adjustment for ethnic group by including a multivariable model adjusting for all covariates included in the original multivariable model apart from ethnic group.

### Patient and public involvement

A community advisory group consisting of migrant women and healthcare professionals was involved in developing the research proposal and provided feedback on the research questions and study design. The community advisory group directly contributed to the decision to include both GP and A&E data, as these were felt particularly relevant to this group of women. The group also offered insights into interpretation of results.

## Results

As shown in Table 1, there were 2,182,280 participants women with singleton pregnancies recorded within MSDS with estimated pregnancy start dates between 1/3/2019-29/2/2024, aged 18-49 years at estimated pregnancy start date. Among participants, 61,140 (2.8%) had a record of interpreter-use, and 2,121,145 (97.2%) did not. The median age of the study population was 31.0 among interpreter-users and 30.9 among non-interpreter-users. Among interpreter-users, 37.5% were of White ethnicity compared to 77.8% of non-interpreter-users. Half (49.7%) of interpreter-users were in the most deprived quintile, compared with 22.7% of non-interpreter-users. Figure 1 shows the reasons for participant exclusions.

**Figure 1.**
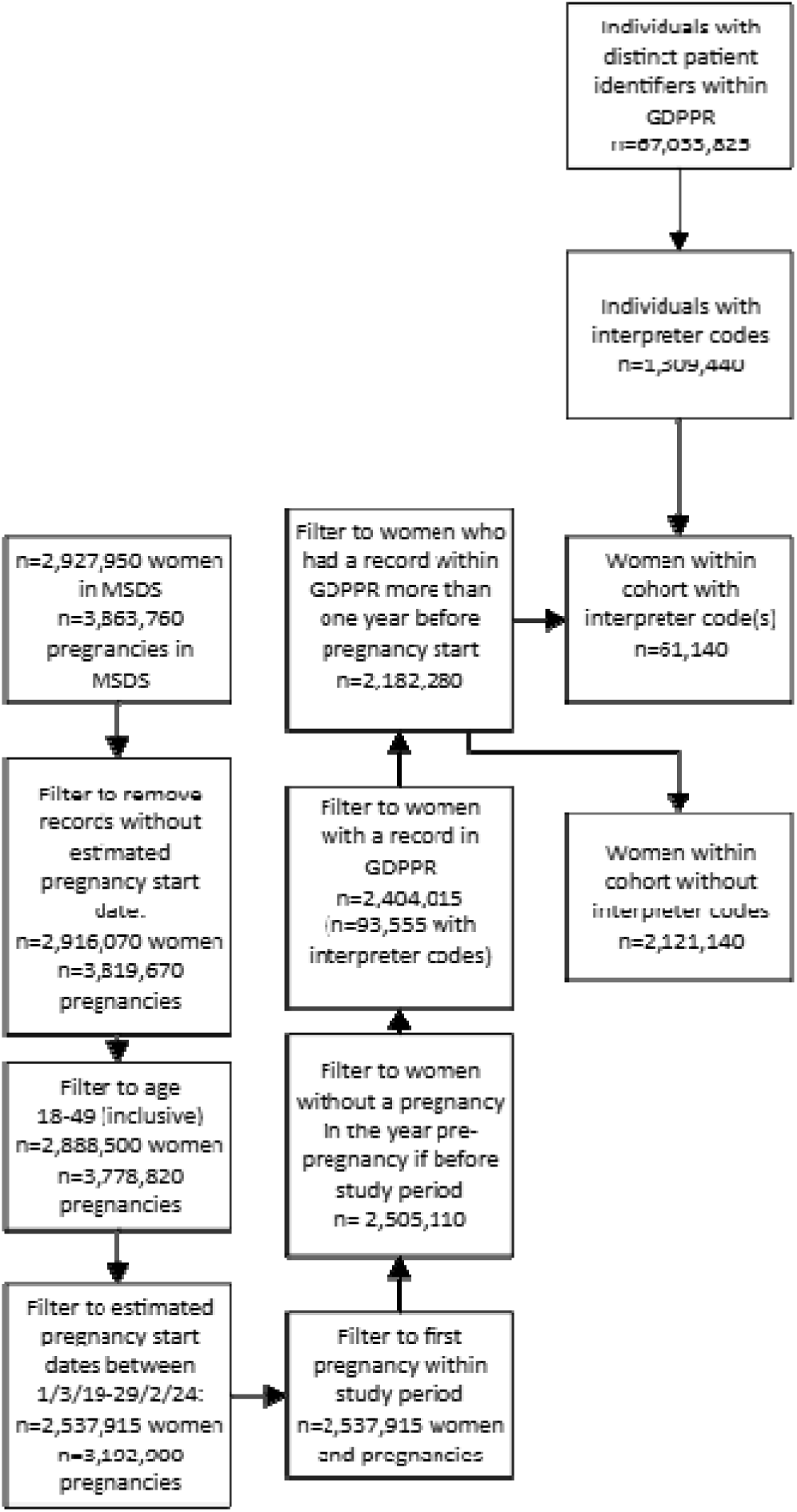
Number of women in the study including exclusions

As shown in Table 2, among both interpreter-users and non-interpreter-users, GP interactions during the year pre-pregnancy were lowest for those whose year pre-pregnancy overlapped with the start of COVID-19 (1^st^ March 2020), and highest for those whose year pre-pregnancy was after the start of COVID-19. GP interactions were lower among interpreter-users than non-interpreter-users both before, during and after the onset of COVID-19.

**Table 2:** Proportion of participants with GP and A&E interactions in the year pre-pregnancy among interpreter-users and non-interpreter-users according to period relative to the onset of COVID-19 (defined as 1/3/2020). Participants consist of women with estimated pregnancy start dates between 1/3/2019-29/2/2024, aged 18-49.

|  | Overall | Interpreter-users | Non-interpreter-users |
| --- | --- | --- | --- |
| **GP interactions in the year pre-pregnancy** |  |  |  |
| **Whole study period** | *n*=2,182,280 | *n*= 61,140 | *n*= 2,121,140 |
| One or more | 1,851,550 (84.8%) | 48,625 (79.5%) | 1,802,925 (85.0%) |
| **Pre-pregnancy year before 1/3/2020** | *N*=547,450 | *N*=15,275 | *N*= 532,175 |
| One or more | 447,450 (81.7%) | 12,280 (80.4%) | 435,165 (81.8%) |
| **Pre-pregnancy year overlaps with 1/3/2020** | *N*= 518,810 | *N*=14,160 | *N*= 504,650 |
| One or more | 415,675 (80.1%) | 10,565 (74.6%) | 405,110 (80.3%) |
| **Pre-pregnancy year after 1/3/2020** | *N*= 1,116,020 | *N*= 31,700 | *N*=1,084,320 |
| One or more | 988,430 (88.6%) | 25,780 (81.3%) | 962,650 (88.8%) |
| **A&E attendance in the year pre-pregnancy** |  |  |  |
| **Whole study period** | *n*=2,182,280 | *n*= 61,140 | *n*= 2,121,140 |
| One or more | 563,675 (25.8%) | 17,240 (28.2%) | 546,440 (25.8%) |
| **Pre-pregnancy year before 1/3/2020** | *N*=547,450 | *N*=15,275 | *N*= 532,175 |
| One or more | 149,470 (27.3%) | 4,465 (29.2%) | 145,000 (27.2%) |
| **Pre-pregnancy year overlaps with 1/3/2020** | *N*= 518,810 | *N*=14,160 | *N*= 504,650 |
| One or more | 121,480 (23.4%) | 3,610 (25.5%) | 117,870 (23.4%) |
| **Pre-pregnancy year after 1/3/2020** | *N*= 1,116,020 | *N*= 31,700 | *N*=1,084,320 |
| One or more | 292,730 (26.2%) | 9,160 (28.9%) | 283,570 (26.2%) |

As shown in Table 3, the adjusted odds of interaction with GP in the year pre-pregnancy were 0.74 (95% CI 0.72-0.75) times lower among interpreter-users versus non-interpreter-users during the study period overall. Before COVID-19, the adjusted odds of GP interaction among interpreter-users compared to non-interpreter-users was 0.92 (95% CI 0.88-0.96); this lowered to 0.63 (95% CI 0.61-0.65) for those with pre-pregnancy years after the onset of COVID-19.

**Table 3.**
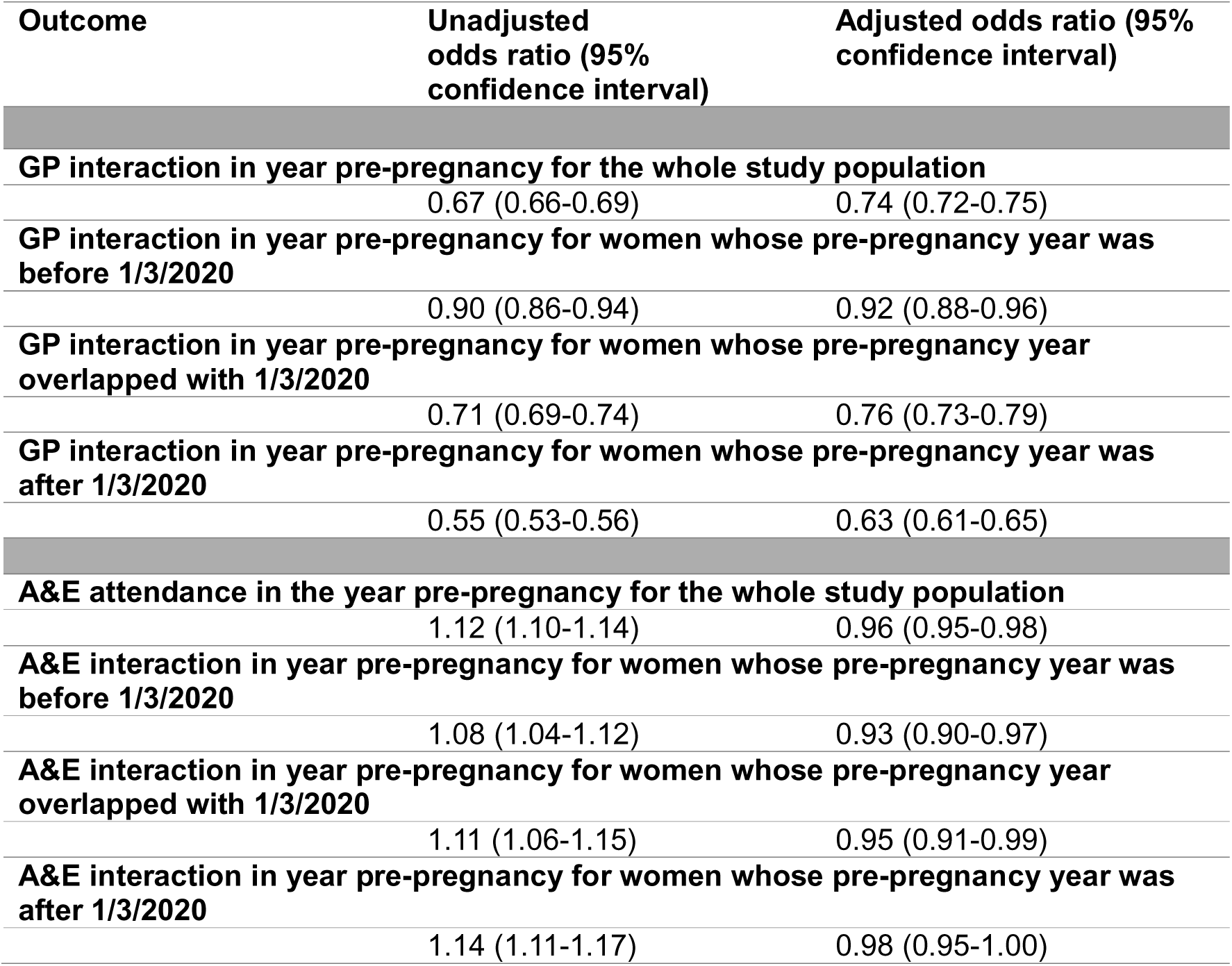
Logistic regression models showing odds of GP interaction and A&E attendance among interpreter-users, with non-interpreter-users as the reference category. Participants consist of women with estimated pregnancy start dates between 1/3/2019-29/2/2024, aged 18-49. *Fully adjusted models adjust for mother’s age (years), ethnic group, geographical region, deprivation quintile and previous pregnancy (binary yes/no). COVID-19 onset defined as 1/3/2020. See Supplementary File 3 for full regression models*.

| Outcome | Unadjusted odds ratio (95% confidence interval) | Adjusted odds ratio (95% confidence interval) |
| --- | --- | --- |
| **GP interaction in year pre-pregnancy for the whole study population** |  |  |
|  | 0.67 (0.66-0.69) | 0.74 (0.72-0.75) |
| **GP interaction in year pre-pregnancy for women whose pre-pregnancy year was before 1/3/2020** |  |  |
|  | 0.90 (0.86-0.94) | 0.92 (0.88-0.96) |
| **GP interaction in year pre-pregnancy for women whose pre-pregnancy year overlapped with 1/3/2020** |  |  |
|  | 0.71 (0.69-0.74) | 0.76 (0.73-0.79) |
| **GP interaction in year pre-pregnancy for women whose pre-pregnancy year was after 1/3/2020** |  |  |
|  | 0.55 (0.53-0.56) | 0.63 (0.61-0.65) |
| **A&E attendance in the year pre-pregnancy for the whole study population** |  |  |
|  | 1.12 (1.10-1.14) | 0.96 (0.95-0.98) |
| **A&E interaction in year pre-pregnancy for women whose pre-pregnancy year was before 1/3/2020** |  |  |
|  | 1.08 (1.04-1.12) | 0.93 (0.90-0.97) |
| **A&E interaction in year pre-pregnancy for women whose pre-pregnancy year overlapped with 1/3/2020** |  |  |
|  | 1.11 (1.06-1.15) | 0.95 (0.91-0.99) |
| **A&E interaction in year pre-pregnancy for women whose pre-pregnancy year was after 1/3/2020** |  |  |
|  | 1.14 (1.11-1.17) | 0.98 (0.95-1.00) |

Overall, A&E interactions were lowest for those whose pre-pregnancy year overlapped with the start of COVID-19 (1^st^ March 2020), and highest for those whose pre-pregnancy year was before the start of COVID-19. A higher proportion of interpreter-users accessed A&E in the year pre-pregnancy than non-interpreter-users during all COVID-19-related time periods.

The unadjusted odds of interaction with A&E in the year pre-pregnancy were 1.12 (95% CI 1.10-1.14) higher among interpreter-users compared with non-interpreter-users, but 0.96 (95% CI 0.95-0.98) times lower among interpreter-users when adjusted for confounding factors. The adjusted odds of A&E interactions did not change substantially between individuals with pre-pregnancy years occurring pre, during and post-COVID-19 onset.

The addition/removal of ethnic group as a covariate did not substantially change odds of GP or A&E interactions (Supplementary File 3), indicating there was no independent contribution of ethnicity on GP/A&E interactions.

See Supplementary File 3 for full regression models including all covariates.

Within the study population, the majority of the interpreter user group (44,245, 72.4%) had their first interpreter code recorded before the year pre-pregnancy (Table 4); 5.2% (n=3,170) had their first interpreter code recorded during the year pre-pregnancy.

**Table 4:** Period of interpreter recording for study participants who have interpreter recorded. Participants consist of women with estimated pregnancy start dates between 1/3/2019-29/2/2024, aged 18-49.

| Period interpreter first recorded within GP record | Women included in the study population<br>Number (%) |
| --- | --- |
| Prior to pre-pregnancy year | 44,245 (72.4%) |
| During year pre-pregnancy | 3,170 (5.2%) |
| After pregnancy start (during pregnancy) | 4,740 (7.8%) |
| After delivery* | 8,985 (14.7%) |

\*For those without a recorded delivery date within the Maternity Services Dataset (MSDS), the last recorded entry for the pregnancy within MSDS was used

## Discussion

### Summary

We have found that the majority of women interact with GP in the year before getting pregnant, and this proportion increased from 85.0% of non-interpreter-users and 79.5% of interpreter-users for pregnancy start dates pre-COVID-19 to 88.8% of non-interpreter-users and 81.3% of interpreter-users for pregnancy start dates post-COVID-19 onset (1/3/2020). Women using interpreters had lower odds of interacting with GP in the year pre-pregnancy compared with non-interpreter-users, and the odds of interaction with GP reduced progressively from pre-COVID-19 (pregnancy start dates occurring before 1/3/2020) to post-COVID-19 (pregnancy start dates occurring after 28/2/2021), suggesting that inequalities between interpreter-users and non-interpreter-users in primary care attendance have increased.

The majority of women included in the study had their first interpreter use recorded before the year pre-pregnancy; but 27.6% did not. This suggests that targeted information about preconception health in languages appropriate for women registered at a practice is challenging because it is not necessarily possible to ascertain which languages women speak until after they have conceived. Ensuring that language preference and need for interpreter is recorded at GP registration is important to ensure that health interventions can be provided in appropriate languages and format.

Most women did not interact with A&E in the year pre-pregnancy, so delivering preconception interventions within A&E would have limited reach compared to primary care. In any case, A&E may be less suited to preconception interventions compared to primary care, given the lack of continuity of care and the focus on urgent versus preventative care.

### Strengths and limitations

This is the first study to explore interactions with GP and A&E in the preconception period since the COVID-19 pandemic, and the first study to explore interactions among women using interpreters. The national linked dataset we used includes primary care, Hospital Episode Statistics and MSDS, providing a unique opportunity to identify women from their first antenatal booking appointment (Hospital Episode Statistics Maternity records do not routinely include antenatal appointments, so largely capture pregnancies that involve a hospital delivery). This means that we were able to include some pregnant women who did not have a successful pregnancy, which previous related studies have not^8^. This means that women who had pregnancy losses after their initial midwife appointment were included; miscarriages occur in 15.3% of recognised pregnancies and these pregnancies may have different preconception risk factors^17^ so are important to consider when undertaking preconception health research.

Baseline characteristics were adjusted for in multivariable analyses but are important to consider when comparing interpreter-users and non-interpreter-users. A real strength of study is that there were very few missing data for ethnic group (0.1%) because ethnicity could be drawn from any of several data sources ^15^; this is particularly important given the well-established evidence on ethnicity and maternal health outcomes^18^. Ethnic group varied between interpreter-users and non-interpreter users: a smaller proportion of interpreter-users were white (37.5%) compared with non-interpreter-users (77.8%). It was not possible to differentiate between ‘White British’ and ‘White other’ ethnic groups. Over two-thirds of interpreter-users (69.0%) had a previous pregnancy, compared with 55.2% of non-interpreter-users. Similarly, 49.7% of interpreter-users were in the most deprived quintile compared with 22.7% of non-interpreter-users. There was geographical variation in proportions of interpreter-users; for example, a larger proportion of interpreter-users (32.1%) were based in London compared with non-interpreter-users (16.0%), which is consistent with patterns of migration^19^.

Although this study provides valuable information about whether women interacted with their GP and A&E in the year pre-pregnancy, we were unable to analyse data on reasons for GP and A&E interactions, although the 500 most frequently occurring codes were reviewed for relevance to GP interactions and are included in Supplementary File 2. We were also unable to identify which languages women spoke, as some codes were limited to simply stating that an interpreter was needed (rather than in what language). Moreover, not all GP codes were available within the data due to the nature of GDPPR having been developed to study COVID-19 and cardiovascular disease^12^; however, the dataset has been used previously to estimate healthcare use^20^ and frequency of GP interactions were similar to those found elsewhere ^8^.

For women with interpreter codes, the code was sourced from a record detailing some interaction in primary care (and did not include free-text records); this means there’s a possibility of self-selection bias because we defined our cases based on interpreter code, which could involve an additional interaction with GP if recorded on a different date to GP registration. People who need an interpreter, but who did not visit the GP so this need was not recorded, are included in the ‘non-interpreter-users’ group and are therefore contributing to an over-estimate of interactions with primary care in the interpreter-users group; however, we still observed that interpreter-users were less likely to interact with GP in the year pre-pregnancy compared with non-interpreter-users, but perhaps this difference between groups is underestimated which would mean inequalities are more substantial than they appear.

Additionally, the inclusion criteria required there to be some GP interaction before the year pre-pregnancy (to ensure they were registered with the GP) so many women using interpreters were excluded from the study population because they had not interacted with the GP before the year preconception (n=32,415, 34.6% of interpreter users pre-exclusions compared to n=189,230, 8.2% of non-interpreter users pre-exclusions). We therefore selected people for inclusion in the study who have been accessing services in some way, again potentially underestimating inequalities since a larger proportion of interpreter-users were excluded for this reason than non-interpreter-users.

MSDS coverage was more limited earlier on during the study^21^, meaning a relatively smaller cohort of women would have been included in earlier years; however, the cohort was still representative of pregnant women in England, and the study was sufficiently powered given the large sample size throughout the study period.

Finally, because we only included the first pregnancy per woman within the study period, it may be assumed that women in later cohorts would be younger; however the median age of the population over the three time periods did not substantially change (median age 30.7 for those whose pregnancy start date was before 1/3/2020, 31.0 for those who pregnancy start date was between 1/3/2020-28/2/2021, and 30.9 for those whose pregnancy start date was after 28/2/2021).

### Comparison with existing literature

This study builds on a previous study using 2017-2018 data exploring pre-pregnancy care in GP in England which found that 86.6% of women had contact with a GP or nurse in the year pre-pregnancy^8^. Our study found that, since COVID-19, this proportion has increased in the general population, indicating that interventions to improve preconception health within primary care could have more reach than ever in the general population, but that women using interpreters are less likely to interact with the GP so may need more targeted approaches to ensure inequalities are not exacerbated.

A previous study exploring migrants’ primary care utilisation before and during COVID-19 in England found that migrants used primary care less than non-migrants, and this difference increased in the first year of the pandemic^22^. Interpreter-users are a subset of migrants that face additional barriers to accessing and using healthcare^23^. Although our study did not examine migrants in particular, there is likely to be significant overlap between interpreter-users and migrants, indicating that both populations may need targeted preconception support outside of primary care.

Rates of A&E attendance were similar in our study compared to national averages (considering that women of childbearing age have higher rates of attendance compared to males)^10^. Although no previous study could be identified which has explored the association between interpreter use and A&E attendance, deprivation is associated with increased attendance^10^ which may explain why odds of A&E attendance in the year preconception were higher among interpreter-users in unadjusted logistic regression models, but not in adjusted models.

Data from the Migration Observatory indicates that 99% of people born in the UK consider English as a first language^24^. Therefore, there is a very small chance that individuals requiring an interpreter were UK-born. This is particularly pertinent given that nearly one in three births in England and Wales were to women born outside the UK in 2023 ^1^, and the recent MBRRACE-UK report highlighting the challenging healthcare environment faced by women with language barriers within maternity^25^.

### Implications for research and practice

We have shown that the majority of women in England interacted with the GP at some point in the year before getting pregnant. Although those using interpreters were less likely to do so, the majority still did, making primary care a promising avenue for addressing preconception health. To address the widening inequalities in accessing primary care in the year preconception, GP practices could actively reach out to people on their lists to obtain information about interpretation needs and pregnancy intentions (where appropriate) to create opportunities to interact. Interventions must be adapted to consider the heterogeneity of the population of interpreter-users. We must also ensure that interventions to improve preconception health are available outside of GP in multiple languages for those who need them including through community groups, health visitors and other routes of access. More research is needed into effective preconception health interventions both within and outside of primary care, particular for women who use interpreters.

## Additional information

### Funding

The British Heart Foundation Data Science Centre (grant No SP/19/3/34678, awarded to Health Data Research (HDR) UK) funded co-development (with NHS England) of the Secure Data Environment service for England, provision of linked datasets, and data management and wrangling support, with additional contributions from the HDR UK Data and Connectivity component of the UK Government Chief Scientific Adviser’s National Core Studies programme to coordinate national COVID-19 priority research. Consortium partner organisations funded the time of contributing data analysts, biostatisticians, epidemiologists, and clinicians.

AS was supported by grants from the British Heart Foundation (RG/18/13/33946: RG/F/23/110103), British Heart Foundation Chair Award (CH/12/2/29428) and by Health Data Research UK, which is funded by the UK Medical Research Council, Engineering and Physical Sciences Research Council, Economic and Social Research Council, Department of Health and Social Care (England), Chief Scientist Office of the Scottish Government Health and Social Care Directorates, Health and Social Care Research and Development Division (Welsh Government), Public Health Agency (Northern Ireland), British Heart Foundation and the Wellcome Trust, National Institute for Health and Care Research (NIHR) Cambridge Biomedical Research Centre (NIHR203312)*, BHF Centre for Research Excellence (RE/18/1/34212 and RE/24/130011), National Institute for Health and Care Research (NIHR303137), Engineering and Physical Sciences Research Council (EP/Y017757/1).

MM is supported by the Medical Research Council [Grant number MR/W01498X/1]. For the purpose of open access, the author has applied a Creative Commons Attribution (CC BY) licence. NP is funded by an NIHR Advanced Fellowship [NIHR305395].

The views expressed in this publication are those of the author(s) and not necessarily those of the NIHR or UKRI.

### Ethical approval

The North East - Newcastle and North Tyneside 2 research ethics committee provided ethical approval for the CVD-COVID-UK/COVID-IMPACT research programme (REC No 20/NE/0161) to access, within secure trusted research environments, unconsented, whole- population, de-identified data from electronic health records collected as part of patients’ routine healthcare.

### Competing interests

None declared

## Acknowledgements

We would like to acknowledge the community advisory group for this project, who provided invaluable support with the design and interpretation of results of the study. We would also like to thank Dr Elena Raffetti for her support in the development of the study and reviewing a draft of the manuscript, and the BHF DSC Health Data Science Team who provided support with protocol development and ad-hoc queries.

This work was carried out with the support of the BHF Data Science Centre led by HDR UK (BHF Grant no. SP/19/3/34678). This study made use of de-identified data held in NHS England’s Secure Data Environment service for England and made available via the BHF Data Science Centre’s CVD-COVID-UK/COVID-IMPACT consortium. This work used data provided by patients and collected by the NHS as part of their care and support. We would also like to acknowledge all data providers who make health relevant data available for research.

## Contributorship statement

MM: conceptualisation, methodology, formal analysis, data curation, writing – original draft, visualisation, funding acquisition. NSC: conceptualisation, methodology, writing – review & editing. LM – methodology, validation, data curation, writing – review & editing. BN – data curation, writing – review & editing. YB – methodology, writing – review & editing. ZW – data curation, writing – review & editing. AS – methodology, writing – review & editing. OO – conceptualisation, methodology, writing – review & editing, supervision. FB – conceptualisation, writing – review & editing, supervision. RA – conceptualisation, methodology, writing – review & editing, supervision. NP - conceptualisation, methodology, writing – review & editing, supervision.

## Data availability

The data used in this study are available in NHS England’s Secure Data Environment (SDE) service for England, but as restrictions apply, they are not publicly available (https://digital.nhs.uk/services/secure-data-environment-service). The CVD-COVID-UK/COVID-IMPACT programme, led by the BHF Data Science Centre (https://bhfdatasciencecentre.org/), received approval to access data in NHS England’s SDE service for England from the Independent Group Advising on the Release of Data (IGARD) (https://digital.nhs.uk/about-nhs-digital/corporate-information-and-documents/independent-group-advising-on-the-release-of-data) via an application made in the Data Access Request Service (DARS) Online system (ref. DARS-NIC-381078-Y9C5K) (https://digital.nhs.uk/services/data-access-request-service-dars/dars-products-and-services). The CVD-COVID-UK/COVID-IMPACT Approvals & Oversight Board (https://bhfdatasciencecentre.org/areas/cvd-covid-uk-covid-impact/) subsequently granted approval to this project (**CCU063_03**) to access the data within NHS England’s SDE service for England. The de-identified data used in this study were made available to accredited researchers only. Those wishing to gain access to the data should contact in the first instance.

## Footnotes

* *We’ve used the terms women/woman, but recognise not all pregnant or birthing people identify as women*.

## Notes

### Competing Interest Statement

The authors have declared no competing interest.

### Clinical Protocols

https://github.com/BHFDSC/CCU063_03

### Author Declarations

The North East - Newcastle and North Tyneside 2 research ethics committee provided ethical approval for the CVD-COVID-UK/COVID-IMPACT research programme (REC No 20/NE/0161) to access within secure trusted research environments unconsented whole-population de-identified data from electronic health records collected as part of patients routine healthcare.

## References

1. Births by parents’ country of birth, England and Wales - Office for National Statistics [Internet]. [cited 2025 Mar 27]. Available from: https://www.ons.gov.uk/peoplepopulationandcommunity/birthsdeathsandmarriages/livebirths/bulletins/parentscountryofbirthenglandandwales/2023

2. MacLellan J, McNiven A, Kenyon S. Provision of interpreting support for cross-cultural communication in UK maternity services: A Freedom of Information request. Int J Nurs Stud Adv. 2024 Jun 1;6:100162.

3. Heslehurst N, Brown H, Pemu A, Coleman H, Rankin J. Perinatal health outcomes and care among asylum seekers and refugees: A systematic review of systematic reviews. BMC Med. 2018;16(1):1–25.

4. Daly M, Kipping RR, Tinner LE, Sanders J, White JW. Preconception exposures and adverse pregnancy, birth and postpartum outcomes: Umbrella review of systematic reviews. Paediatr Perinat Epidemiol. 2022;36(2):288–99.

5. McGranahan M, Augarde E, Schoenaker D, Duncan H, Mann S, Bick D, et al. Preconception health among migrant women in England: A cross-sectional analysis of maternity services data 2018–2019. J Migr Health. 2024 Jan 1;10:100250.

6. Perinatal confidential enquiry | MBRRACE-UK. The care of recent migrant women with language barriers who have experienced a stillbirth or neonatal death. [Internet]. [cited 2025 Mar 27]. Available from: https://timms.le.ac.uk/mbrrace-uk-perinatal-mortality/confidential-enquiries/confidential-enquiry-migrant-women.html

7. Withanage NN, Botfield JR, Srinivasan S, Black KI, Mazza D. Effectiveness of preconception interventions in primary care: a systematic review. British Journal of General Practice [Internet]. 2022 Dec 1 [cited 2025 Mar 28];72(725):e865–72. Available from: https://bjgp.org/content/72/725/e865

8. Li Y, Kurinczuk JJ, Alderdice F, Quigley MA, Rivero-Arias O, Sanders J, et al. Pre-pregnancy care in general practice in England: cross-sectional observational study using administrative routine health data. BMC Public Health 2025 25:1 [Internet]. 2025 Mar 22 [cited 2025 Mar 28];25(1):1–12. Available from: https://bmcpublichealth.biomedcentral.com/articles/10.1186/s12889-025-21728-1

9. Zhao T, Meacock R, Sutton M. Drivers of primary care appointment volumes before and after the COVID-19 pandemic: a longitudinal study. BMC Health Serv Res [Internet]. 2025 Dec 1 [cited 2025 May 7];25(1):1–10. Available from: https://bmchealthservres.biomedcentral.com/articles/10.1186/s12913-025-12488-0

10. Inequalities in Accident and Emergency department attendance, England - Office for National Statistics [Internet]. [cited 2025 Mar 28]. Available from: https://cy.ons.gov.uk/peoplepopulationandcommunity/healthandsocialcare/healthcaresystem/articles/inequalitiesinaccidentandemergencydepartmentattendanceengland/march2021tomarch2022

11. GPES data for pandemic planning and research (COVID-19) (GDPPR) - NHS England Digital [Internet]. [cited 2025 May 22]. Available from: https://digital.nhs.uk/coronavirus/gpes-data-for-pandemic-planning-and-research

12. Wood A, Denholm R, Hollings S, Cooper J, Ip S, Walker V, et al. Linked electronic health records for research on a nationwide cohort of more than 54 million people in England: Data resource. The BMJ [Internet]. 2021 Apr 7 [cited 2025 Jun 9];373. Available from: https://pubmed.ncbi.nlm.nih.gov/33827854/

13. GDPPR | BHF DSC HDS Documentation [Internet]. [cited 2025 May 19]. Available from: https://bhfdsc.github.io/documentation/docs/dataset_insights/gdppr

14. Pathak N, Zhang CX, Boukari Y, Burns R, Mathur R, Gonzalez-Izquierdo A, et al. Development and Validation of a Primary Care Electronic Health Record Phenotype to Study Migration and Health in the UK. Int J Environ Res Public Health [Internet]. 2021 Dec 17;18(24):13304. Available from: https://www.mdpi.com/1660-4601/18/24/13304

15. Methodology | BHF DSC HDS Documentation [Internet]. [cited 2025 May 11]. Available from: https://bhfdsc.github.io/documentation/curated_assets/kpcs/methodology

16. Vissandjee B, Desmeules M, Cao Z, Abdool S, Kazanjian A. Integrating Ethnicity and Migration As Determinants of Canadian Women’s Health. BMC Womens Health [Internet]. 2004 Aug 1 [cited 2025 May 19];4(Suppl 1):1–11. Available from: https://link.springer.com/articles/10.1186/1472-6874-4-S1-S32

17. Quenby S, Gallos ID, Dhillon-Smith RK, Podesek M, Stephenson MD, Fisher J, et al. Miscarriage matters: the epidemiological, physical, psychological, and economic costs of early pregnancy loss. The Lancet [Internet]. 2021 May 1 [cited 2025 Jun 25];397(10285):1658–67. Available from: https://www.thelancet.com/action/showFullText?pii=S0140673621006826

18. Felker A, Patel R, Kotnis R, Kenyon S, Knight M, on behalf of MBRRACE-UK. Saving Lives, Improving Mothers’ Care. Lessons learned to inform maternity care from the UK and Ireland Confidential Enquiries into Maternal Deaths and Morbidity 2020-22 [Internet]. Oxford; 2024 [cited 2025 Mar 28]. Available from: www.hqip.org.uk/national-programmes.

19. The changing picture of long-term international migration, England and Wales - Office for National Statistics [Internet]. [cited 2025 May 7]. Available from: https://www.ons.gov.uk/peoplepopulationandcommunity/populationandmigration/internationalmigration/articles/thechangingpictureoflongterminternationalmigrationenglandandwales/census2021

20. Mu Y, Dashtban A, Mizani MA, Tomlinson C, Mohamed M, Ashworth M, et al. Healthcare utilisation of 282,080 individuals with long COVID over two years: a multiple matched control, longitudinal cohort analysis. J R Soc Med [Internet]. 2024 Nov 1 [cited 2025 Mar 28];117(11). Available from: https://pubmed.ncbi.nlm.nih.gov/39603265/

21. NHS Maternity Statistics, England, 2023-24 - NHS England Digital [Internet]. [cited 2025 Jun 27]. Available from: https://digital.nhs.uk/data-and-information/publications/statistical/nhs-maternity-statistics/2023-24

22. Zhang CX, Boukari Y, Pathak N, Mathur R, Katikireddi SV, Patel P, et al. Migrants’ primary care utilisation before and during the COVID-19 pandemic in England: An interrupted time series analysis. The Lancet Regional Health - Europe [Internet]. 2022 Sep 1 [cited 2025 Mar 28];20:100455. Available from: https://www.thelancet.com/action/showFullText?pii=S2666776222001491

23. Pandey M, Maina RG, Amoyaw J, Li Y, Kamrul R, Michaels CR, et al. Impacts of English language proficiency on healthcare access, use, and outcomes among immigrants: a qualitative study. BMC Health Serv Res [Internet]. 2021 Dec 1 [cited 2025 May 7];21(1):1–13. Available from: https://link.springer.com/articles/10.1186/s12913-021-06750-4

24. English language use and proficiency of migrants in the UK - Migration Observatory - The Migration Observatory [Internet]. [cited 2025 Jun 27]. Available from: https://migrationobservatory.ox.ac.uk/resources/briefings/english-language-use-and-proficiency-of-migrants-in-the-uk/

25. Perinatal confidential enquiry | MBRRACE-UK The care of recent migrant women with language barriers who have experienced a stillbirth or neonatal death [Internet]. [cited 2025 Jun 27]. Available from: https://timms.le.ac.uk/mbrrace-uk-perinatal-mortality/confidential-enquiries/confidential-enquiry-migrant-women.html

