## Supplementary File 1 for "Pre-pregnancy primary care and accident and emergency interactions among interpreter-users versus non-interpreter-users: a retrospective cohort study"

**Pregnancy start date**

Pregnancy start date (i.e. conception) was estimated from fields within MSDS (the Maternity Services Dataset). One field within MSDS, ‘eddagreed’, is derived from other fields. We prioritised the source of pregnancy start date as follows:

1. Eddagreed Method 02, minus 280 days
2. Eddagreed Method 03, minus 280 days
3. Procedure date for dating ultrasound scan minus gestational age at dating ultrasound scan
4. Eddagreed Method 01 minus 280 days
5. Last menstrual period date
6. Eddagreed Method 04 minus 280 days
7. Appointment date for formal antenatal booking minus gestational age at booking

*‘Eddagreed’ Method of calculation*

Method 01 – Last Menstrual Period (LMP) date stated by mother

Method 02 – LMP date confirmed by ultrasound scan in pregnancy

Method 03 – Ultrasound scan in pregnancy dating measurements

Method 04 – Clinical assessment

**Gestation variables considered plausible (days)**

- Minimum plausible gestation = 28
- Maximum plausible gestation = 315
- Minimum livebirths = 0
- Maximum livebirths = 24
- Minimum stillbirths = 0
- Maximum stillbirths = 24
- Minimum early loss = 0
- Maximum early loss = 30

**How we recorded previous pregnancy**

Previous pregnancy was based on the MSDS variables ‘previous still births’, ‘previous live births’ and ‘previous losses less than 24 weeks’; if an individual had one or more of any of these three variables, they were recorded as having a previous pregnancy.

**Codes removed from top 500 interactions because not considered a GP interaction**

*# 1240781000000106: Severe acute respiratory syndrome coronavirus 2 vaccination invitation short message service text message sent (situation)*

*# 1109911000000100: Excepted from cervical screening quality indicators - no response to three invitations (finding)*

*# 1300561000000107: High risk category for developing complication from coronavirus disease 19 caused by severe acute respiratory syndrome coronavirus 2 infection (finding)*

*# 171152003: Ca cervix screening - not wanted (situation)*

*# 185731000: Asthma monitoring call first letter (procedure)*

*# 956951000000104: Pertussis vaccination in pregnancy (procedure)*

*# 1109921000000106: Quality and Outcomes Framework quality indicator-related care invitation (procedure)*

*# 185732007: Asthma monitoring call second letter (procedure)*

*# 391156007: Medication review without patient (procedure)*

*# 185736005: Asthma monitoring call telephone invite (procedure)*

*# 1300591000000101: Low risk category for developing complication from coronavirus disease 19 caused by severe acute respiratory syndrome coronavirus 2 infection (finding)*

*# 1300571000000100: Moderate risk category for developing complication from coronavirus disease 19 caused by severe acute respiratory syndrome coronavirus 2 infection (finding)*

*# 1324721000000108: Severe acute respiratory syndrome coronavirus 2 vaccination dose declined (situation)*

*# 1090701000000104: National Health Service Diabetes Prevention Programme invitation (procedure)*

*# 783401000000101: Stop smoking invitation first short message service text message (procedure)*

*# 783381000000101: Stop smoking invitation short message service text message (procedure)*

*# 976631000000101: White: English or Welsh or Scottish or Northern Irish or British - England and Wales ethnic category 2011 census (finding)*

*# 976691000000100: White: any other White background - England and Wales ethnic category 2011 census (finding)*

*# 976791000000107: Asian or Asian British: Indian - England and Wales ethnic category 2011 census (finding)*

*# 976811000000108: Asian or Asian British: Pakistani - England and Wales ethnic category 2011 census (finding)*

*# 976871000000103: Asian or Asian British: any other Asian background - England and Wales ethnic category 2011 census (finding)*

*# 976891000000104: Black or African or Caribbean or Black British: African - England and Wales ethnic category 2011 census (finding)*

*# 18167009: Black African (ethnic group)*

*# 414481008: Indian (racial group)*

*# 494131000000105: White British - ethnic category 2001 census (finding)*

*# 92391000000108: British or mixed British - ethnic category 2001 census (finding)*

*# 315236000: White British (ethnic group)*

*# 92411000000108: Other White background - ethnic category 2001 census (finding)*

*# 110751000000108: Indian or British Indian - ethnic category 2001 census (finding)*

*# 92461000000105: Pakistani or British Pakistani - ethnic category 2001 census (finding)*

*# 92531000000104: Ethnic category not stated - 2001 census (finding)*

*# 92491000000104: African - ethnic category 2001 census (finding)*

*# 185984009: White - ethnic group (ethnic group)*

*# 92481000000101: Other Asian background - ethnic category 2001 census (finding)*

*# 110761000000106: English - ethnic category 2001 census (finding)*

*# 92471000000103: Bangladeshi or British Bangladeshi - ethnic category 2001 census (finding)*

*# 92521000000101: Other - ethnic category 2001 census (finding)*

*# 94041000000106: Other White European or European unspecified or Mixed European - ethnic category 2001 census (finding)*

*# 92451000000107: Other Mixed background - ethnic category 2001 census (finding)*

*# 92511000000107: Chinese - ethnic category 2001 census (finding)*

*# 92421000000102: White and Black Caribbean - ethnic category 2001 census (finding)*

*# 94051000000109: Other White or White unspecified - ethnic category 2001 census (finding)*

*# 88941000000100: Polish - ethnic category 2001 census (finding)*

*# 92431000000100: White and Black African - ethnic category 2001 census (finding)*

*# 107691000000105: Caribbean - ethnic category 2001 census (finding)*

*# 94151000000105: Any other group - ethnic category 2001 census (finding)*

*# 976871000000103: Asian or Asian British: any other Asian background - England and Wales ethnic category 2011 census (finding)*

*# 976971000000106: Other ethnic group: any other ethnic group - England and Wales ethnic category 2011 census (finding)*

*# 92441000000109: White and Asian - ethnic category 2001 census (finding)*

*# 89001000000105: Arab - ethnic category 2001 census (finding)*

*# 186002003: Pakistani (ethnic group)*
