## Supplementary File 3 for "Pre-pregnancy primary care and accident and emergency interactions among interpreter-users versus non-interpreter-users: a retrospective cohort study"

Supplementary File 3: Full regression models

| Univariable analysis for outcome GP interaction preconception whole study period | | | | |
| --- | --- | --- | --- | --- |
| term | Odds ratio | P value | 95% confidence interval - lower bound | 95% confidence interval - upper bound |
| Interpreter use | 0.67 | <0.001 | 0.66 | 0.69 |

| Univariable analysis for outcome GP interaction preconception for women whose pre-pregnancy year was before 1/3/2020 | | | | |
| --- | --- | --- | --- | --- |
|  |  | P value | 95% confidence interval - lower bound | 95% confidence interval - upper bound |
| Interpreter use | 0.90 | <0.001 | 0.86 | 0.94 |

| Univariable analysis for outcome GP interaction preconception for women whose pre-pregnancy year overlapped with 1/3/2020 | | | | |
| --- | --- | --- | --- | --- |
| term | Odds ratio | P value | 95% confidence interval - lower bound | 95% confidence interval - upper bound |
| Interpreter use | 0.71 | <0.001 | 0.69 | 0.74 |

| Univariable analysis for outcome GP interaction preconception for women whose pre-pregnancy year was after 1/3/2020 | | | | |
| --- | --- | --- | --- | --- |
| term | Odds ratio | P value | 95% confidence interval - lower bound | 95% confidence interval - upper bound |
| Interpreter use | 0.55 | <0.001 | 0.53 | 0.56 |

| Multivariable regression model for outcome: GP interaction preconception during whole study period | | | |  |
| --- | --- | --- | --- | --- |
|  | Odds ratio | P value | 95% confidence interval - lower bound | 95% confidence interval - upper bound |
| Interpreter use | 0.77 | <0.001 | 0.75 | 0.78 |
| Age (years) | 1.01 | <0.001 | 1.01 | 1.01 |
| Ethnicity |  |  |  |  |
| White | Reference |  |  |  |
| Black, Black British, Caribbean or African | 0.78 | <0.001 | 0.76 | 0.79 |
| Asian or Asian British | 0.91 | <0.001 | 0.90 | 0.92 |
| Mixed or multiple ethnic groups | 0.89 | <0.001 | 0.87 | 0.91 |
| Other ethnic group | 0.74 | <0.001 | 0.72 | 0.76 |
| Deprivation quintile |  |  |  |  |
| IMD 1 - most deprived | Reference |  |  |  |
| IMD 2 | 1.03 | <0.001 | 1.02 | 1.05 |
| IMD 3 | 1.06 | <0.001 | 1.05 | 1.07 |
| IMD 4 | 1.06 | <0.001 | 1.04 | 1.07 |
| IMD 5 - least deprived | 1.06 | <0.001 | 1.04 | 1.07 |
| Region |  |  |  |  |
| London | Reference | | | |
| South East | 1.22 | <0.001 | 1.20 | 1.24 |
| South West | 1.36 | <0.001 | 1.34 | 1.39 |
| East of England | 1.35 | <0.001 | 1.33 | 1.37 |
| East Midlands | 1.34 | <0.001 | 1.31 | 1.36 |
| West Midlands | 1.22 | <0.001 | 1.20 | 1.23 |
| Yorkshire and The Humber | 1.50 | <0.001 | 1.47 | 1.52 |
| North East | 1.58 | <0.001 | 1.54 | 1.61 |
| North West | 1.31 | <0.001 | 1.29 | 1.33 |
| Previous pregnancy | 1.14 | <0.001 | 1.13 | 1.15 |

| Multivariable regression model for outcome GP interaction preconception without ethnicity whole study period | | | | |
| --- | --- | --- | --- | --- |
|  | Odds ratio | P value | 95% confidence interval - lower bound | 95% confidence interval - upper bound |
| Interpreter use | 0.70 | <0.001 | 0.68 | 0.71 |
| Age (years) | 1.01 | <0.001 | 1.01 | 1.01 |
| Deprivation quintile |  |  |  |  |
| IMD 1 - most deprived | Reference | | |  |
| IMD 2 | 1.05 | <0.001 | 1.03 | 1.06 |
| IMD 3 | 1.09 | <0.001 | 1.07 | 1.10 |
| IMD 4 | 1.09 | <0.001 | 1.07 | 1.10 |
| IMD 5 - least deprived | 1.09 | <0.001 | 1.08 | 1.11 |
| Region |  |  |  |  |
| London | Reference |  |  |  |
| South East | 1.28 | <0.001 | 1.26 | 1.29 |
| South West | 1.45 | <0.001 | 1.42 | 1.47 |
| East of England | 1.41 | <0.001 | 1.39 | 1.44 |
| East Midlands | 1.40 | <0.001 | 1.38 | 1.43 |
| West Midlands | 1.26 | <0.001 | 1.24 | 1.28 |
| Yorkshire and The Humber | 1.58 | <0.001 | 1.56 | 1.61 |
| North East | 1.69 | <0.001 | 1.65 | 1.72 |
| North West | 1.38 | <0.001 | 1.36 | 1.40 |
| Previous pregnancy | 1.14 | <0.001 | 1.13 | 1.15 |

| Multivariable regression model for outcome: GP interaction preconception for women whose pre-pregnancy year was before 1/3/2020 | | | |  |
| --- | --- | --- | --- | --- |
|  | Odds ratio | P value | 95% confidence interval - lower bound | 95% confidence interval - upper bound |
| Interpreter use | 0.92 | <0.001 | 0.88 | 0.96 |
| Age (years) | 1.00 | 0.026 | 1.00 | 1.00 |
| Ethnicity |  |  |  |  |
| White | Reference |  |  |  |
| Black, Black British, Caribbean or African | 0.87 | <0.001 | 0.84 | 0.90 |
| Asian or Asian British | 0.91 | <0.001 | 0.89 | 0.94 |
| Mixed or multiple ethnic groups | 0.90 | <0.001 | 0.86 | 0.94 |
| Other ethnic group | 0.77 | <0.001 | 0.74 | 0.81 |
| Deprivation quintile |  |  |  |  |
| IMD 1 - most deprived | Reference |  |  |  |
| IMD 2 | 0.97 | 0.003 | 0.94 | 0.99 |
| IMD 3 | 0.98 | 0.206 | 0.96 | 1.01 |
| IMD 4 | 0.97 | 0.006 | 0.94 | 0.99 |
| IMD 5 - least deprived | 0.97 | 0.042 | 0.95 | 1.00 |
| Region |  |  |  |  |
| London | Reference | | | |
| South East | 1.16 | <0.001 | 1.13 | 1.19 |
| South West | 1.26 | <0.001 | 1.22 | 1.30 |
| East of England | 1.28 | <0.001 | 1.24 | 1.32 |
| East Midlands | 1.32 | <0.001 | 1.28 | 1.37 |
| West Midlands | 1.21 | <0.001 | 1.18 | 1.25 |
| Yorkshire and The Humber | 1.41 | <0.001 | 1.37 | 1.46 |
| North East | 1.32 | <0.001 | 1.27 | 1.38 |
| North West | 1.24 | <0.001 | 1.21 | 1.28 |
| Previous pregnancy | 1.37 | <0.001 | 1.35 | 1.40 |

| Multivariable regression model for outcome GP interaction preconception without ethnicity for women whose pre-pregnancy year was before 1/3/2020 | | | | |
| --- | --- | --- | --- | --- |
|  | Odds ratio | P value | 95% confidence interval - lower bound | 95% confidence interval - upper bound |
| Interpreter use | 0.88 | <0.001 | 0.84 | 0.92 |
| Age (years) | 1.00 | 0.239 | 1.00 | 1.00 |
| Deprivation quintile |  |  |  |  |
| IMD 1 - most deprived | Reference | | |  |
| IMD 2 | 0.97 | 0.022 | 0.95 | 1.00 |
| IMD 3 | 1.00 | 0.898 | 0.98 | 1.03 |
| IMD 4 | 0.99 | 0.254 | 0.96 | 1.01 |
| IMD 5 - least deprived | 1.00 | 0.761 | 0.97 | 1.02 |
| Region |  |  |  |  |
| London | Reference |  |  |  |
| South East | 1.20 | <0.001 | 1.17 | 1.23 |
| South West | 1.32 | <0.001 | 1.28 | 1.36 |
| East of England | 1.33 | <0.001 | 1.29 | 1.37 |
| East Midlands | 1.37 | <0.001 | 1.33 | 1.42 |
| West Midlands | 1.25 | <0.001 | 1.21 | 1.28 |
| Yorkshire and The Humber | 1.47 | <0.001 | 1.43 | 1.52 |
| North East | 1.39 | <0.001 | 1.33 | 1.45 |
| North West | 1.29 | <0.001 | 1.25 | 1.33 |
| Previous pregnancy | 1.37 | <0.001 | 1.35 | 1.39 |

| Multivariable regression model for outcome: GP interaction preconception for women whose pre-pregnancy year overlapped with 1/3/2020 | | | |  |
| --- | --- | --- | --- | --- |
|  | Odds ratio | P value | 95% confidence interval - lower bound | 95% confidence interval - upper bound |
| Interpreter use | 0.76 | <0.001 | 0.73 | 0.79 |
| Age (years) | 1.00 | 0.725 | 1.00 | 1.00 |
| Ethnicity |  |  |  |  |
| White | Reference |  |  |  |
| Black, Black British, Caribbean or African | 0.82 | <0.001 | 0.79 | 0.85 |
| Asian or Asian British | 0.86 | <0.001 | 0.84 | 0.88 |
| Mixed or multiple ethnic groups | 0.92 | <0.001 | 0.88 | 0.96 |
| Other ethnic group | 0.78 | <0.001 | 0.75 | 0.82 |
| Deprivation quintile |  |  |  |  |
| IMD 1 - most deprived | Reference |  |  |  |
| IMD 2 | 1.01 | 0.577 | 0.98 | 1.03 |
| IMD 3 | 1.00 | 0.986 | 0.98 | 1.02 |
| IMD 4 | 1.01 | 0.334 | 0.99 | 1.04 |
| IMD 5 - least deprived | 0.98 | 0.212 | 0.96 | 1.01 |
| Region |  |  |  |  |
| London | Reference | | | |
| South East | 1.19 | <0.001 | 1.16 | 1.22 |
| South West | 1.36 | <0.001 | 1.32 | 1.40 |
| East of England | 1.41 | <0.001 | 1.37 | 1.45 |
| East Midlands | 1.31 | <0.001 | 1.27 | 1.36 |
| West Midlands | 1.20 | <0.001 | 1.17 | 1.24 |
| Yorkshire and The Humber | 1.56 | <0.001 | 1.51 | 1.61 |
| North East | 1.51 | <0.001 | 1.45 | 1.58 |
| North West | 1.34 | <0.001 | 1.31 | 1.38 |
| Previous pregnancy | 1.25 | <0.001 | 1.23 | 1.27 |

| Multivariable regression model for outcome GP interaction preconception without ethnicity for women whose pre-pregnancy year overlapped with 1/3/2020 | | | | |
| --- | --- | --- | --- | --- |
|  | Odds ratio | P value | 95% confidence interval - lower bound | 95% confidence interval - upper bound |
| Interpreter use | 0.72 | <0.001 | 0.69 | 0.75 |
| Age (years) | 1.00 | 0.038 | 1.00 | 1.00 |
| Deprivation quintile |  |  |  |  |
| IMD 1 - most deprived | Reference | | |  |
| IMD 2 | 1.02 | 0.093 | 1.00 | 1.04 |
| IMD 3 | 1.03 | 0.035 | 1.00 | 1.05 |
| IMD 4 | 1.04 | 0.001 | 1.02 | 1.07 |
| IMD 5 - least deprived | 1.02 | 0.167 | 0.99 | 1.04 |
| Region |  |  |  |  |
| London | Reference |  |  |  |
| South East | 1.24 | <0.001 | 1.21 | 1.28 |
| South West | 1.44 | <0.001 | 1.40 | 1.49 |
| East of England | 1.47 | <0.001 | 1.43 | 1.52 |
| East Midlands | 1.38 | <0.001 | 1.33 | 1.42 |
| West Midlands | 1.24 | <0.001 | 1.21 | 1.28 |
| Yorkshire and The Humber | 1.64 | <0.001 | 1.59 | 1.69 |
| North East | 1.62 | <0.001 | 1.55 | 1.68 |
| North West | 1.41 | <0.001 | 1.37 | 1.45 |
| Previous pregnancy | 1.24 | <0.001 | 1.22 | 1.26 |

| Multivariable regression model for outcome: GP interaction preconception for women whose pre-pregnancy year was after 1/3/2020 | | | |  |
| --- | --- | --- | --- | --- |
|  | Odds ratio | P value | 95% confidence interval - lower bound | 95% confidence interval - upper bound |
| Interpreter use | 0.63 | <0.001 | 0.61 | 0.65 |
| Age (years) | 1.02 | <0.001 | 1.02 | 1.02 |
| Ethnicity |  |  |  |  |
| White | Reference |  |  |  |
| Black, Black British, Caribbean or African | 0.67 | <0.001 | 0.65 | 0.69 |
| Asian or Asian British | 0.90 | <0.001 | 0.88 | 0.91 |
| Mixed or multiple ethnic groups | 0.82 | <0.001 | 0.79 | 0.85 |
| Other ethnic group | 0.66 | <0.001 | 0.64 | 0.68 |
| Deprivation quintile |  |  |  |  |
| IMD 1 - most deprived | Reference |  |  |  |
| IMD 2 | 1.10 | <0.001 | 1.08 | 1.12 |
| IMD 3 | 1.17 | <0.001 | 1.15 | 1.19 |
| IMD 4 | 1.19 | <0.001 | 1.17 | 1.21 |
| IMD 5 - least deprived | 1.26 | <0.001 | 1.24 | 1.29 |
| Region |  |  |  |  |
| London | Reference | | | |
| South East | 1.30 | <0.001 | 1.27 | 1.33 |
| South West | 1.49 | <0.001 | 1.45 | 1.53 |
| East of England | 1.37 | <0.001 | 1.33 | 1.40 |
| East Midlands | 1.35 | <0.001 | 1.32 | 1.39 |
| West Midlands | 1.25 | <0.001 | 1.22 | 1.28 |
| Yorkshire and The Humber | 1.55 | <0.001 | 1.51 | 1.59 |
| North East | 1.74 | <0.001 | 1.68 | 1.81 |
| North West | 1.34 | <0.001 | 1.31 | 1.37 |
| Previous pregnancy | 1.20 | <0.001 | 1.19 | 1.22 |

| Multivariable regression model for outcome GP interaction preconception without ethnicity for women whose pre-pregnancy year was after 1/3/2020 | | | | |
| --- | --- | --- | --- | --- |
|  | Odds ratio | P value | 95% confidence interval - lower bound | 95% confidence interval - upper bound |
| Interpreter use | 0.59 | <0.001 | 0.57 | 0.61 |
| Age (years) | 1.02 | <0.001 | 1.02 | 1.02 |
| Deprivation quintile |  |  |  |  |
| IMD 1 - most deprived | Reference | | |  |
| IMD 2 | 1.12 | <0.001 | 1.10 | 1.14 |
| IMD 3 | 1.21 | <0.001 | 1.19 | 1.24 |
| IMD 4 | 1.24 | <0.001 | 1.22 | 1.27 |
| IMD 5 - least deprived | 1.33 | <0.001 | 1.30 | 1.35 |
| Region |  |  |  |  |
| London | Reference |  |  |  |
| South East | 1.39 | <0.001 | 1.36 | 1.42 |
| South West | 1.62 | <0.001 | 1.58 | 1.67 |
| East of England | 1.46 | <0.001 | 1.43 | 1.50 |
| East Midlands | 1.45 | <0.001 | 1.41 | 1.49 |
| West Midlands | 1.32 | <0.001 | 1.29 | 1.35 |
| Yorkshire and The Humber | 1.68 | <0.001 | 1.64 | 1.72 |
| North East | 1.92 | <0.001 | 1.86 | 1.99 |
| North West | 1.45 | <0.001 | 1.42 | 1.48 |
| Previous pregnancy | 1.20 | <0.001 | 1.18 | 1.21 |

| Univariable analysis for outcome A&E interaction preconception whole study period | | | | |
| --- | --- | --- | --- | --- |
| term | Odds ratio | P value | 95% confidence interval - lower bound | 95% confidence interval - upper bound |
| Interpreter use | 1.12 | <0.001 | 1.10 | 1.14 |

| Univariable analysis for outcome A&E interaction preconception for women whose pre-pregnancy year was before 1/3/2020 | | | | |
| --- | --- | --- | --- | --- |
|  |  | P value | 95% confidence interval - lower bound | 95% confidence interval - upper bound |
| Interpreter use | 1.08 | <0.001 | 1.04 | 1.12 |

| Univariable analysis for outcome A&E interaction preconception for women whose pre-pregnancy year overlapped with 1/3/2020 | | | | |
| --- | --- | --- | --- | --- |
| term | Odds ratio | P value | 95% confidence interval - lower bound | 95% confidence interval - upper bound |
| Interpreter use | 1.11 | <0.001 | 1.06 | 1.15 |

| Univariable analysis for outcome A&E interaction preconception for women whose pre-pregnancy year was after 1/3/2020 | | | | |
| --- | --- | --- | --- | --- |
| term | Odds ratio | P value | 95% confidence interval - lower bound | 95% confidence interval - upper bound |
| Interpreter use | 1.14 | <0.001 | 1.11 | 1.17 |

| Multivariable regression model for outcome: A&E interaction preconception during whole study period | | | |  |
| --- | --- | --- | --- | --- |
|  | Odds ratio | P value | 95% confidence interval - lower bound | 95% confidence interval - upper bound |
| Interpreter use | 0.96 | <0.001 | 0.95 | 0.98 |
| Age (years) | 0.94 | <0.001 | 0.94 | 0.94 |
| Ethnicity |  |  |  |  |
| White | Reference |  |  |  |
| Black, Black British, Caribbean or African | 1.17 | <0.001 | 1.15 | 1.19 |
| Asian or Asian British | 0.89 | <0.001 | 0.88 | 0.90 |
| Mixed or multiple ethnic groups | 1.12 | <0.001 | 1.10 | 1.14 |
| Other ethnic group | 0.98 | 0.041 | 0.96 | 1.00 |
| Deprivation quintile |  |  |  |  |
| IMD 1 - most deprived | Reference |  |  |  |
| IMD 2 | 0.87 | <0.001 | 0.86 | 0.88 |
| IMD 3 | 0.79 | <0.001 | 0.78 | 0.79 |
| IMD 4 | 0.72 | <0.001 | 0.71 | 0.73 |
| IMD 5 - least deprived | 0.64 | <0.001 | 0.64 | 0.65 |
| Region |  |  |  |  |
| London | Reference | | | |
| South East | 0.98 | <0.001 | 0.96 | 0.99 |
| South West | 0.77 | <0.001 | 0.76 | 0.78 |
| East of England | 0.90 | <0.001 | 0.88 | 0.91 |
| East Midlands | 0.91 | <0.001 | 0.90 | 0.92 |
| West Midlands | 0.93 | <0.001 | 0.92 | 0.94 |
| Yorkshire and The Humber | 0.95 | <0.001 | 0.93 | 0.96 |
| North East | 1.11 | <0.001 | 1.09 | 1.12 |
| North West | 1.06 | <0.001 | 1.04 | 1.07 |
| Previous pregnancy | 1.43 | <0.001 | 1.42 | 1.44 |

| Multivariable regression model for outcome A&E interaction preconception without ethnicity whole study period | | | | |
| --- | --- | --- | --- | --- |
|  | Odds ratio | P value | 95% confidence interval - lower bound | 95% confidence interval - upper bound |
| Interpreter use | 0.95 | <0.001 | 0.93 | 0.97 |
| Age (years) | 0.94 | <0.001 | 0.94 | 0.94 |
| Deprivation quintile |  |  |  |  |
| IMD 1 - most deprived | Reference | | |  |
| IMD 2 | 0.87 | <0.001 | 0.86 | 0.88 |
| IMD 3 | 0.78 | <0.001 | 0.78 | 0.79 |
| IMD 4 | 0.72 | <0.001 | 0.71 | 0.72 |
| IMD 5 - least deprived | 0.64 | <0.001 | 0.64 | 0.65 |
| Region |  |  |  |  |
| London | Reference |  |  |  |
| South East | 0.97 | <0.001 | 0.96 | 0.98 |
| South West | 0.77 | <0.001 | 0.75 | 0.78 |
| East of England | 0.89 | <0.001 | 0.88 | 0.90 |
| East Midlands | 0.91 | <0.001 | 0.89 | 0.92 |
| West Midlands | 0.92 | <0.001 | 0.91 | 0.93 |
| Yorkshire and The Humber | 0.94 | <0.001 | 0.93 | 0.95 |
| North East | 1.10 | <0.001 | 1.08 | 1.12 |
| North West | 1.05 | <0.001 | 1.03 | 1.06 |
| Previous pregnancy | 1.43 | <0.001 | 1.42 | 1.44 |

| Multivariable regression model for outcome: A&E interaction preconception for women whose pre-pregnancy year was before 1/3/2020 | | | |  |
| --- | --- | --- | --- | --- |
|  | Odds ratio | P value | 95% confidence interval - lower bound | 95% confidence interval - upper bound |
| Interpreter use | 0.93 | 0.001 | 0.90 | 0.97 |
| Age (years) | 0.95 | <0.001 | 0.95 | 0.95 |
| Ethnicity |  |  |  |  |
| White | Reference |  |  |  |
| Black, Black British, Caribbean or African | 1.19 | <0.001 | 1.15 | 1.22 |
| Asian or Asian British | 0.90 | <0.001 | 0.88 | 0.92 |
| Mixed or multiple ethnic groups | 1.13 | <0.001 | 1.09 | 1.18 |
| Other ethnic group | 0.96 | 0.042 | 0.91 | 1.00 |
| Deprivation quintile |  |  |  |  |
| IMD 1 - most deprived | Reference |  |  |  |
| IMD 2 | 0.87 | <0.001 | 0.86 | 0.89 |
| IMD 3 | 0.78 | <0.001 | 0.77 | 0.80 |
| IMD 4 | 0.71 | <0.001 | 0.70 | 0.73 |
| IMD 5 - least deprived | 0.64 | <0.001 | 0.63 | 0.66 |
| Region |  |  |  |  |
| London | Reference | | | |
| South East | 0.92 | <0.001 | 0.90 | 0.95 |
| South West | 0.76 | <0.001 | 0.74 | 0.78 |
| East of England | 0.79 | <0.001 | 0.77 | 0.81 |
| East Midlands | 0.89 | <0.001 | 0.86 | 0.92 |
| West Midlands | 0.88 | <0.001 | 0.86 | 0.91 |
| Yorkshire and The Humber | 0.90 | <0.001 | 0.87 | 0.92 |
| North East | 1.09 | <0.001 | 1.05 | 1.13 |
| North West | 1.03 | 0.010 | 1.01 | 1.06 |
| Previous pregnancy | 1.30 | <0.001 | 1.29 | 1.32 |

| Multivariable regression model for outcome A&E interaction preconception without ethnicity for women whose pre-pregnancy year was before 1/3/2020 | | | | |
| --- | --- | --- | --- | --- |
|  | Odds ratio | P value | 95% confidence interval - lower bound | 95% confidence interval - upper bound |
| Interpreter use | 0.92 | <0.001 | 0.88 | 0.95 |
| Age (years) | 0.95 | <0.001 | 0.94 | 0.95 |
| Deprivation quintile |  |  |  |  |
| IMD 1 - most deprived | Reference | | |  |
| IMD 2 | 0.87 | <0.001 | 0.85 | 0.89 |
| IMD 3 | 0.78 | <0.001 | 0.76 | 0.80 |
| IMD 4 | 0.71 | <0.001 | 0.70 | 0.73 |
| IMD 5 - least deprived | 0.64 | <0.001 | 0.63 | 0.65 |
| Region |  |  |  |  |
| London | Reference |  |  |  |
| South East | 0.92 | <0.001 | 0.90 | 0.94 |
| South West | 0.76 | <0.001 | 0.74 | 0.78 |
| East of England | 0.79 | <0.001 | 0.76 | 0.81 |
| East Midlands | 0.88 | <0.001 | 0.86 | 0.91 |
| West Midlands | 0.87 | <0.001 | 0.85 | 0.90 |
| Yorkshire and The Humber | 0.89 | <0.001 | 0.86 | 0.91 |
| North East | 1.08 | <0.001 | 1.04 | 1.12 |
| North West | 1.02 | 0.051 | 1.00 | 1.05 |
| Previous pregnancy | 1.31 | <0.001 | 1.29 | 1.33 |

| Multivariable regression model for outcome: A&E interaction preconception for women whose pre-pregnancy year overlapped with 1/3/2020 | | | |  |
| --- | --- | --- | --- | --- |
|  | Odds ratio | P value | 95% confidence interval - lower bound | 95% confidence interval - upper bound |
| Interpreter use | 0.95 | 0.027 | 0.91 | 0.99 |
| Age (years) | 0.94 | 0.000 | 0.94 | 0.94 |
| Ethnicity |  |  |  |  |
| White | Reference |  |  |  |
| Black, Black British, Caribbean or African | 1.16 | <0.001 | 1.12 | 1.20 |
| Asian or Asian British | 0.86 | <0.001 | 0.84 | 0.88 |
| Mixed or multiple ethnic groups | 1.13 | <0.001 | 1.08 | 1.18 |
| Other ethnic group | 0.97 | 0.265 | 0.93 | 1.02 |
| Deprivation quintile |  |  |  |  |
| IMD 1 - most deprived | Reference |  |  |  |
| IMD 2 | 0.86 | <0.001 | 0.84 | 0.87 |
| IMD 3 | 0.76 | <0.001 | 0.74 | 0.78 |
| IMD 4 | 0.70 | <0.001 | 0.69 | 0.72 |
| IMD 5 - least deprived | 0.62 | <0.001 | 0.60 | 0.63 |
| Region |  |  |  |  |
| London | Reference | | | |
| South East | 0.95 | <0.001 | 0.93 | 0.97 |
| South West | 0.75 | <0.001 | 0.73 | 0.78 |
| East of England | 0.87 | <0.001 | 0.84 | 0.89 |
| East Midlands | 0.87 | <0.001 | 0.84 | 0.89 |
| West Midlands | 0.87 | <0.001 | 0.85 | 0.90 |
| Yorkshire and The Humber | 0.93 | <0.001 | 0.90 | 0.95 |
| North East | 1.07 | <0.001 | 1.03 | 1.11 |
| North West | 1.02 | 0.097 | 1.00 | 1.05 |
| Previous pregnancy | 1.33 | <0.001 | 1.31 | 1.35 |

| Multivariable regression model for outcome A&E interaction preconception without ethnicity for women whose pre-pregnancy year overlapped with 1/3/2020 | | | | |
| --- | --- | --- | --- | --- |
|  | Odds ratio | P value | 95% confidence interval - lower bound | 95% confidence interval - upper bound |
| Interpreter use | 0.93 | 0.001 | 0.89 | 0.97 |
| Age (years) | 0.94 | 0.000 | 0.94 | 0.94 |
| Deprivation quintile |  |  |  |  |
| IMD 1 - most deprived | Reference | | |  |
| IMD 2 | 0.85 | 0.000 | 0.84 | 0.87 |
| IMD 3 | 0.76 | 0.000 | 0.74 | 0.78 |
| IMD 4 | 0.70 | 0.000 | 0.69 | 0.72 |
| IMD 5 - least deprived | 0.62 | 0.000 | 0.60 | 0.63 |
| Region |  |  |  |  |
| London | Reference |  |  |  |
| South East | 0.95 | <0.001 | 0.93 | 0.97 |
| South West | 0.76 | <0.001 | 0.74 | 0.78 |
| East of England | 0.87 | <0.001 | 0.84 | 0.89 |
| East Midlands | 0.87 | <0.001 | 0.84 | 0.89 |
| West Midlands | 0.87 | <0.001 | 0.84 | 0.89 |
| Yorkshire and The Humber | 0.92 | <0.001 | 0.89 | 0.95 |
| North East | 1.07 | <0.001 | 1.03 | 1.11 |
| North West | 1.02 | 0.140 | 0.99 | 1.05 |
| Previous pregnancy | 1.33 | <0.001 | 1.31 | 1.35 |

| Multivariable regression model for outcome: A&E interaction preconception for women whose pre-pregnancy year was after 1/3/2020 | | | |  |
| --- | --- | --- | --- | --- |
|  | Odds ratio | P value | 95% confidence interval - lower bound | 95% confidence interval - upper bound |
| Interpreter use | 0.98 | 0.132 | 0.95 | 1.01 |
| Age (years) | 0.94 | 0.000 | 0.94 | 0.94 |
| Ethnicity |  |  |  |  |
| White | Reference |  |  |  |
| Black, Black British, Caribbean or African | 1.16 | 0.000 | 1.14 | 1.19 |
| Asian or Asian British | 0.90 | 0.000 | 0.89 | 0.91 |
| Mixed or multiple ethnic groups | 1.11 | 0.000 | 1.08 | 1.14 |
| Other ethnic group | 0.99 | 0.299 | 0.96 | 1.01 |
| Deprivation quintile |  |  |  |  |
| IMD 1 - most deprived | Reference |  |  |  |
| IMD 2 | 0.88 | 0.000 | 0.87 | 0.89 |
| IMD 3 | 0.80 | 0.000 | 0.79 | 0.81 |
| IMD 4 | 0.73 | 0.000 | 0.72 | 0.74 |
| IMD 5 - least deprived | 0.66 | 0.000 | 0.65 | 0.67 |
| Region |  |  |  |  |
| London | Reference | | | |
| South East | 1.01 | 0.290 | 0.99 | 1.03 |
| South West | 0.78 | <0.001 | 0.76 | 0.80 |
| East of England | 0.96 | <0.001 | 0.94 | 0.98 |
| East Midlands | 0.94 | <0.001 | 0.92 | 0.96 |
| West Midlands | 0.97 | <0.001 | 0.95 | 0.98 |
| Yorkshire and The Humber | 0.97 | 0.006 | 0.96 | 0.99 |
| North East | 1.12 | <0.001 | 1.10 | 1.15 |
| North West | 1.08 | <0.001 | 1.06 | 1.10 |
| Previous pregnancy | 1.56 | <0.001 | 1.54 | 1.57 |

| Multivariable regression model for outcome A&E interaction preconception without ethnicity for women whose pre-pregnancy year was after 1/3/2020 | | | | |
| --- | --- | --- | --- | --- |
|  | Odds ratio | P value | 95% confidence interval - lower bound | 95% confidence interval - upper bound |
| Interpreter use | 0.97 | 0.013 | 0.94 | 0.99 |
| Age (years) | 0.94 | <0.001 | 0.94 | 0.94 |
| Deprivation quintile |  |  |  |  |
| IMD 1 - most deprived | Reference | | |  |
| IMD 2 | 0.88 | <0.001 | 0.86 | 0.89 |
| IMD 3 | 0.80 | <0.001 | 0.79 | 0.81 |
| IMD 4 | 0.73 | <0.001 | 0.72 | 0.74 |
| IMD 5 - least deprived | 0.66 | <0.001 | 0.65 | 0.67 |
| Region |  |  |  |  |
| London | Reference |  |  |  |
| South East | 1.00 | 0.608 | 0.99 | 1.02 |
| South West | 0.78 | <0.001 | 0.76 | 0.79 |
| East of England | 0.96 | <0.001 | 0.94 | 0.97 |
| East Midlands | 0.94 | <0.001 | 0.92 | 0.95 |
| West Midlands | 0.96 | <0.001 | 0.94 | 0.97 |
| Yorkshire and The Humber | 0.97 | <0.001 | 0.95 | 0.98 |
| North East | 1.12 | <0.001 | 1.09 | 1.14 |
| North West | 1.07 | <0.001 | 1.06 | 1.09 |
| Previous pregnancy | 1.56 | <0.001 | 1.55 | 1.58 |
